# How to better communicate exponential growth of infectious diseases

**DOI:** 10.1101/2020.06.12.20129114

**Authors:** Martin Schonger, Daniela Sele

## Abstract

Exponential growth bias is the phenomenon that humans underestimate exponential growth. In the context of infectious diseases, this bias may lead to failure to understand the effectiveness of non-pharmaceutical interventions (NPIs). Communicating the same scenario in different ways (framing) has been found to have a large impact on people’s evaluations and behavior in the contexts of social behavior, risk taking and health care. We find that framing matters for people’s assessment of the benefits of NPIs. In two commonly used frames, most subjects in our experiment drastically underestimate the number of cases NPIs avoid. Framing growth in terms of doubling times, rather than growth rates, reduces bias. When the scenario is framed in terms of time gained, rather than cases avoided, the median subject assesses the benefit of NPIs correctly. These findings suggest changes that public health authorities can adopt to better communicate the exponential spread of infectious diseases.

## INTRODUCTION

Non-pharmaceutical interventions (NPIs) like social distancing, wearing masks, (precautionary) quarantines, school closures or dialling down economic life have been found to contain the spread of COVID-19 (*1-3*). Absent options like vaccines or treatments, such behavioral adaptations may need to be sustained for many months. However, while their costs are directly felt, their benefit is abstract: they slow the prospective exponential growth of the infectious disease. Public support for and adherence to such measures depends on correct assessment of their benefits, in particular by opinion and community leaders. Previous research (*4-14*) has shown that people underestimate exponential growth. For financial contexts, empirical research has shown that biased perception of exponential growth causally affects real-world behaviour (*11*), and for the infectious disease context, recent lab evidence suggests the same (*4*).

The assessment of exponential disease spread, and of the benefit of decreasing the underlying growth rate, may depend on the way information is communicated. Indeed, regarding other decision problems in social behaviour (*15,16*), risk taking (*17,20*) and health care (*21-24*), different ways of communicating identical information about a scenario, i.e. different frames, have been found to alter people’s perception and evaluation of available choices. In this study, we experimentally investigate whether different frames can facilitate understanding of exponential growth and of the benefit of NPIs.

In the experiment we present subjects with a hypothetical scenario, in which a country can slow the spread of an infectious disease by adopting NPIs. The country has 974 cases of the infectious disease. Without these mitigation measures, the number of cases grows by 26% a day, to about 1 million cases in 30 days. NPIs would reduce the daily growth rate to 9% a day, resulting in about 13,000 cases in 30 days. Or, looking at the benefit of the mitigation measures from a different perspective, the country would gain about 50 days until 1 million cases are reached.

Subjects are asked their beliefs concerning three questions: what the benefit of the mitigation measures is (‘mitigation question’), how much the disease spreads if no mitigation measures are taken (‘high exponential growth question’) and by how much it spreads if mitigation measures are taken (‘low exponential growth question’).

The scenario and the three questions are presented to subjects in one of four frames. Each subject is randomly assigned to one frame. Frames vary along two dimensions, r vs. d and C vs. T. Dimension r vs. d concerns the way in which the growth of the disease is communicated: either in terms of the daily growth rate (r), or in the equivalent doubling time in days (d). Dimension C vs. T changes the perspective on the benefit of the mitigation measures: either in terms of the cases avoided within 30 days (about 985,000 in C) or in terms of the time gained until 1 million cases are reached (about 50 days in T). This results in four frames, C-r, C-d, T-r, and T-d, which are given in table 1.

**Table 1.** Experimental stimuli in the four different frames.

| **All frames**<br>In a country, there are 974 cases of an infectious disease. |  |
| --- | --- |
| **Frame C-r**<br><u>Mitigation question:</u><br>Mitigation measures would reduce the daily growth rate from 26% to 9%. How many cases could be avoided in 30 days?<br>(986,330 cases)<br><u>High [low] exponential growth question:</u><br>The number of infected people grows by 26% [9%] per day. How many people will be infected in 30 days?<br>(999,253 [12,923] cases) | **Frame C-d**<br><u>Mitigation question:</u><br>Mitigation measures would lengthen the doubling time from 3 days to 8 days. How many cases could be avoided in 30 days?<br>(984,271 cases)<br><u>High [low] exponential growth question:</u><br>The number of infected people doubles every 3 [8] days. How many people will be infected in 30 days?<br>(997,376 [13,105] cases) |
| **Frame T-r**<br><u>Mitigation question:</u><br>Mitigation measures would reduce the daily growth rate from 26% to 9%. How much time could be gained until 1 million cases are reached?<br>( 50.46 days)<br><u>High [low] exponential growth question:</u><br>The number of infected people grows by 26% [9%] per day. How long will it take until 1,000,000 people are infected?<br>( 30.00 [80.46] days) | **Frame T-d**<br><u>Mitigation question:</u><br>Mitigation measures would lengthen the doubling time from 3 days to 8 days. How much time could be gained until 1 million cases are reached?<br>( 50.02 days)<br><u>High [low] exponential growth question:</u><br>The number of infected people doubles every 3 [8] days. How long will it take until 1,000,000 people are infected?<br>( 30.01 [80.03] days) |
| **Table 1 \| Experimental stimuli in the four different frames.** |  |

Fig. 1 illustrates the underlying system. The parameters of the questions are set such that for the high growth exponential question the correct answer in frames C-r/C-d is about equal to the number of cases given in frames T-r/T-d, and, conversely, the correct answer in frames T-r/T-d is about equal to the amount of time given in frames C-r/C-d. The mitigation measures either reduce the number of cases in the country (C-r/C-d), or buy time for the country (T-r/T-d).

**Fig 1.**
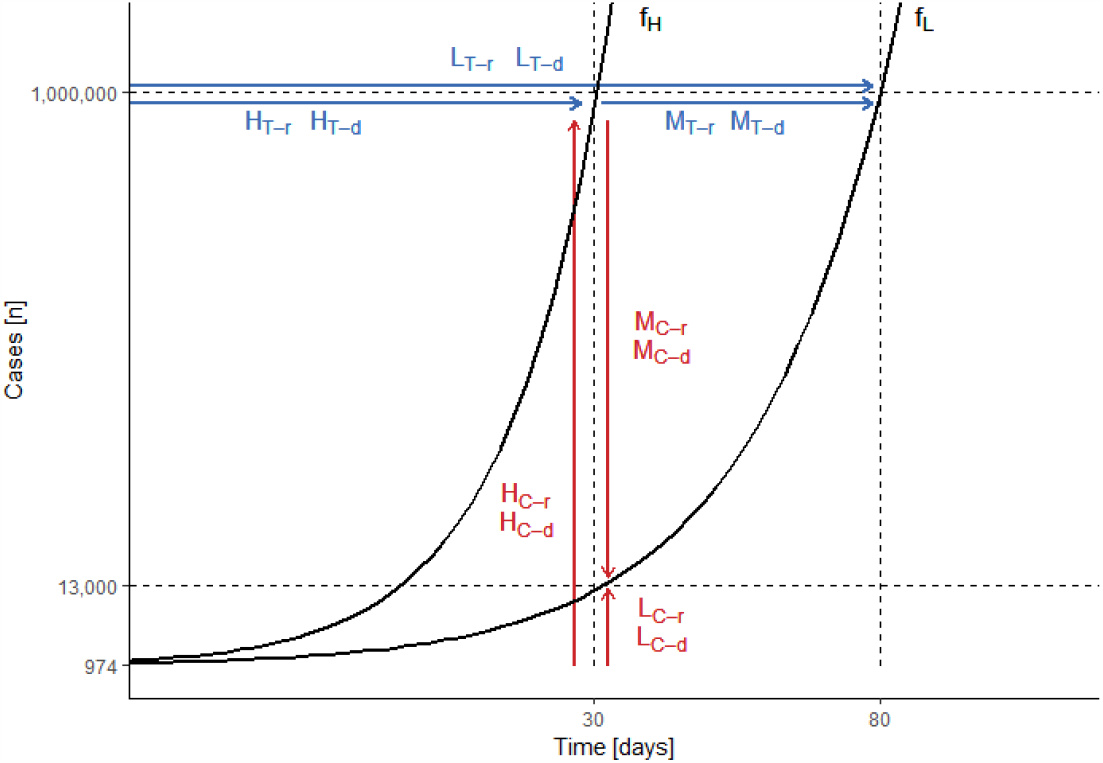
Schematic of questions, with exponential functions *f*_H_(26% per day/doubling time of 3 days) and *f*_L_ (9% per day/doubling time of 8 days). For each frame F, M_F_ depicts the mitigation, H_F_ the high exponential growth, L_F_ the low exponential growth question.

Hence, in all four frames subjects are given the same exponential function, but while in frames C-r/C-d they are asked for cases as a function of time, an exponential problem, in frames T-r/T-d they are asked for time as a function of cases, a logarithmic problem. Anthropological evidence finds that people naturally employ logarithmic scales (*25,26*). However, this does not imply that they find logarithmic problems intuitive.

A subject exhibits exponential growth bias if she underestimates exponential growth. In frames C-r/C-d, this means underestimating the number of cases which result after a given time. In frames T-r/T-d, this means overestimating the amount of time until a given number of cases is reached. In line with this, we define mitigation bias as underestimating the benefit of decelerating the exponential spread of the disease. In frames C-r/C-d, this means underestimating the number of cases avoided due to the mitigation measures, in frames T-r/T-d it means underestimating the number of days gained due to the measures.

The study was conducted online on March 25 and 26, 2020. At this time, all educational institutions and non-essential shops in Switzerland were closed due to SARS-CoV-2. Subjects were students in non-STEM fields at Swiss universities. To get at subjects’ intuitive perception of exponential growth, rather than their calculating skills, subjects were requested to refrain from using calculators or other tools. The order of questions was randomized. For further study details, see Methods. In total, there were 459 subjects, 116 each in frames C-r, T-r and T-d, and 111 in frame C-d.

## RESULTS AND DISCUSSION

### Subjects underestimate the benefits of NPIs, but framing can eliminate the bias

Fig. 2 gives the cumulative distribution functions of subjects’ answers, with the solid vertical lines indicating the correct answers and the shaded areas indicating beliefs with mitigation/exponential growth bias. For the mitigation question, in frames C-r/C-d (fig. 2a), NPIs avoid about 985,000 cases. In both frames, most subjects underestimate these benefits: 94% of subjects in frame C-r and 87% of subjects in frame C-d exhibit mitigation bias. These shares are not statistically significantly different at the 99%-level. The median answer is 8,600 cases avoided in frame C-r, and 82,000 cases avoided in frame C-d (for further quantiles see extended data table 1). Hence, the median answer in the frame using doubling times exhibits less bias than the median answer in the frame using growth rates (*p* < 10^−5^). For frames T-r/T-d, where mitigation measures buy the country about 50 days, consider fig. 2b: 44% of subjects in frame T-r and 36% of subjects in frame T-d believe that fewer days are gained, i.e. exhibit mitigation bias. The fraction of subjects exhibiting mitigation bias is not statistically significantly different between the two frames at the 99%-level. The median assessment of the benefit of the mitigation measures is 60 days in frame T-r, and 50 days in frame T-d. The median answers are not statistically significantly different between the two frames at the 99%-level. Frames C-r/C-d employ the exponential perspective, and frames T-r/T-d employ the logarithmic perspective. Therefore the median answers are not comparable, but one can compare these frames according to the fraction of biased subjects. In frame C-r, 50 percentage points more subjects are biased than in frame T-r (*p* < 10^−14^). In frame C-d, 51 percentage points more subjects are biased than in frame T-d (*p* < 10^−13^).

**Fig 2.**
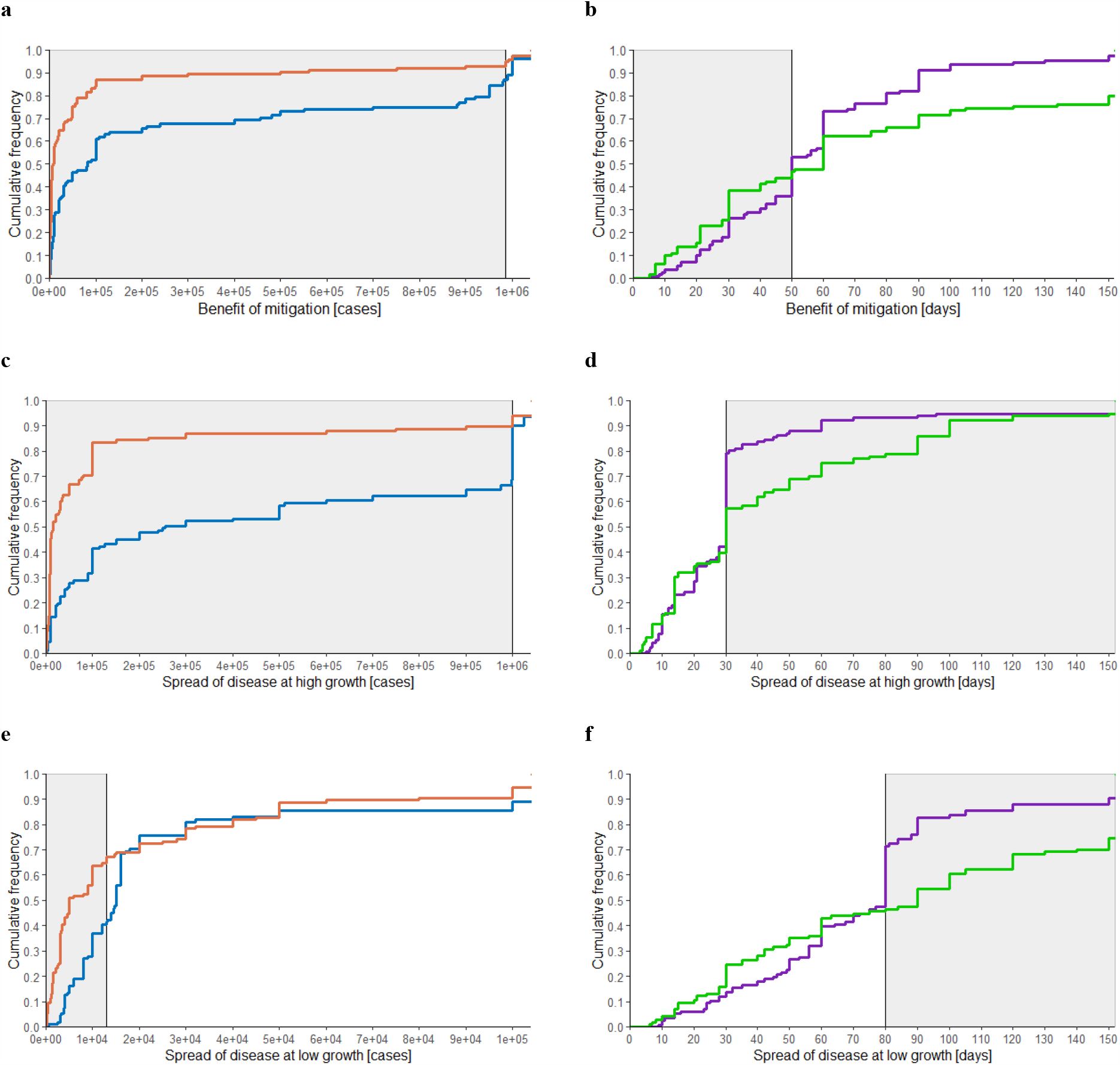
Effect of framing on bias, **a-b**, cumulative distribution function (CDF) of answers to the mitigation question, for **a**, frame C-r (orange line, n=114)) and frame C-d (blue line, n=102), **b**, frame T-r (green line, n=109) and frame T-d (purple line, n=111), **c-d**, CDF of answers to the high exponential growth question, **c**, frame C-r (n=115) and C-d (n=111), **d**, frame T-r (n=109) and T-d (n=111), **e-f**, CDF of answers to the low exponential growth question, **e**, frame C-r (n=116) and C-d (n=111), **f**, frame T-r (n=113) and T-d (n=116). Solid vertical line indicates correct answer. Shaded area indicates beliefs which reveal mitigation /exponential growth bias. Axes are capped.

### Framing matters for the perception of the spread of infectious disease

If the disease spreads at the high growth rate, there will be about 1 million cases in 30 days (fig. 2c). 90% of subjects in frame C-r and 67% of subjects in frame C-d underestimate this, i.e. exhibit exponential growth bias. The median answer is 15,000 cases in frame C-r, and 256,000 cases in frame C-d. Framing the scenario using doubling times facilitates understanding: the share of subjects who exhibit the bias is lower (*p* < 10^−4^), and the median answer in that frame is closer to the correct amount (*p* < 10^−3^). Turning to frames T-r/T-d, the median answer in these frames coincides with the correct value of 30 days. 42% of subjects in frame T-r, and 21% of subjects in frame T-d believe it takes longer than that to reach 1 million cases, i.e. exhibit exponential growth bias. Hence, the share of participants exhibiting exponential growth bias is lower when doubling times are used (*p* < 10^−3^). Comparing the exponential and logarithmic perspectives, in frame C-r 48 percentage points more subjects are biased than in frame T-r (*p* < 10^−13^), and in frame C-d, 46 percentage points more subjects are biased than in frame T-d (*p* < 10^−11^).

If the disease spreads at the low growth rate, there will be about 13,000 cases after 30 days (fig. 2e). In frame C-r, the median answer is 5,000 cases, and 65% of subjects exhibit exponential growth bias. In frame C-d, which uses doubling times, the median answer is 15,000 cases and only a minority of subjects, 41%, exhibit exponential growth bias. The median answer in the frame using doubling times is closer to the correct amount than the median answer in the frame using growth rates. For frames T-r/T-d, the correct answer is about 80 days (fig. 2f). The median answer is 90 days in frame T-r, and 80 days in frame T-d. 54% of subjects in frame T-r, and 28% of subjects in frame T-d believe it takes longer to reach a million cases. Hence, we again find that the share of participants exhibiting exponential growth bias is lower when doubling times are used (*p* < 10^−4^). Comparing A vs. B, in frame C-r, 11 percentage points more subjects are biased than in frame T-r (*p* = 0.056), and in frame C-d, 13 percentage points more subjects are biased than in frame T-d (*p* = 0.03).

### The frame using doubling time and the logarithmic perspective has the fewest biased subjects

Table 2 compares any two frames, giving the difference in the share of biased subjects. The picture that emerges is that some ways to communicate exponential growth are better than others. First, compare communicating growth in terms of doubling times to communicating growth in terms of growth rates. In any comparison that differs only along this dimension, the share of biased subjects is smaller when doubling times are used. Second, in any comparison that differs only in whether the logarithmic perspective rather than the exponential perspective is prompted, the share of biased subjects is smaller with the logarithmic perspective. That is, when asked for time rather than for cases, fewer subjects exhibit mitigation bias and exponential growth bias. Hence, in all questions, the fraction of subjects exhibiting bias is lower in frame T-d than in all other frames.

**Table 2.** Difference in share of biased subjects across frames. The 95%-confidence intervals are given in parentheses. The p-value of the one-sided hypothesis test is given.

| Mitigation question |  |  |  |
| --- | --- | --- | --- |
|  | C-d | T-r | T-d |
| C-r | -7%<br>(-15%, 1%)<br>$p = 0.07$ | -50%<br>(-60%, -40%)<br>$p < 10^{-14}$ | -58%<br>(-68%, -48%)<br>$p < 10^{-16}$ |
| C-d | - | -43%<br>(-54%, -32%)<br>$p < 10^{-10}$ | -51%<br>(-62%, -40%)<br>$p < 10^{-13}$ |
| T-r | - | - | -8%<br>(-21%, 5%)<br>$p = 0.14$ |
| High exponential growth question |  |  |  |
|  | C-d | T-r | T-d |
| C-r | -24%<br>(-34%, -13%)<br>$p < 10^{-4}$ | -48%<br>(-59%, -37%)<br>$p < 10^{-13}$ | -70%<br>(-79%, -61%)<br>$p < 10^{-16}$ |
| C-d | - | -24%<br>(-37%, -12%)<br>$p < 10^{-3}$ | -46%<br>(-57%, -35%)<br>$p < 10^{-11}$ |
| T-r | - | - | -22%<br>(-34%, -10%)<br>$p < 10^{-3}$ |
| Low exponential growth question |  |  |  |
|  | C-d | T-r | T-d |
| C-r | -23%<br>(-36%, -11%)<br>$p < 10^{-3}$ | -11%<br>(-24%, 1%)<br>$p = 0.056$ | -36%<br>(-48%, -24%)<br>$p < 10^{-7}$ |
| C-d | - | 12%<br>(-0.1%, 25%)<br>$p = 0.46$ | -13%<br>(-25%, -0.1%)<br>$p = 0.03$ |
| T-r | - | - | -25%<br>(-37%, -13%)<br>$p < 10^{-4}$ |

### Mitigation bias is larger than what exponential growth bias would predict

In the following, we examine to what extent mitigation bias can be accounted for by exponential growth bias. The correct answer to the mitigation question is the difference between the correct answers to the two exponential growth questions. Hence, to relate the mitigation bias found in frames C-r and C-d to exponential growth bias, we compare a subject’s answer in the mitigation question to the difference between her answers to the high and low exponential growth questions (fig. 3). For this exercise, we restrict attention to positive data points and those subjects to whom the mitigation question was displayed prior to the exponential growth questions. For 14% of subjects in frame C-r and 5% of subjects in frame C-d, the answer to the mitigation question is exactly equal to the difference in their answers to the exponential growth questions (for subjects who see the mitigation question after the exponential growth questions, this occurs more frequently, see extended data figure 1). 66% of subjects in frame C-r and 76% of subjects in frame C-d give an answer to the mitigation question which is smaller than the difference in their answers to the exponential growth questions. Hence, for these subjects mitigation bias is larger than one would expect based on their answers to the exponential growth questions. The null hypothesis that mitigation bias is not larger than exponential growth bias is rejected for both frame C-r (*p* < 0.05) and frame C-d (*p* < 10^−4^).

**Fig 3.**
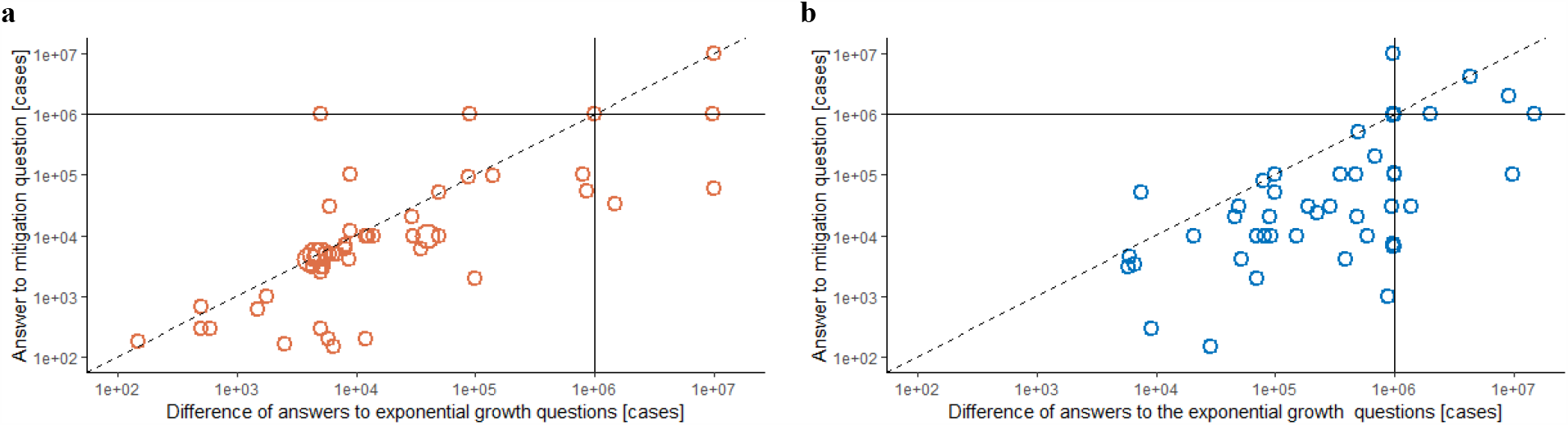
Mitigation bias and exponential growth bias. **a**, answers to the mitigation question plotted against the difference in answers to the exponential growth questions for frame C-r (n=54), **b**, same plot for frame C-d (n=50). Solid lines indicate the correct answer to the mitigation question resp. the difference between the correct answers to the exponential growth questions (about 1 million cases avoided). For observations on the dashed line, mitigation bias can be fully explained by exponential growth bias. Multiple identical answers are displayed by larger circles. Only subjects to whom the mitigation question was displayed prior to the exponential growth questions. Data points with non-positive values excluded, one large outlier in C-d not shown.

### Being aware of exponential growth bias does not prevent it

The phenomenon of exponential growth bias has been discussed for centuries (*27,28*) and has received renewed attention in the press in the context of the coronavirus pandemic (*29,30*). To investigate subjects’ awareness of exponential growth bias, we ask subjects what they believe about other subjects’ answers to the high exponential growth question. 83% of subjects in frame C-r and 91% of subjects in frame C-d believe that others underestimate or strongly underestimate the number of cases. In both frames T-r and T-d, 66% of subjects believe that others overestimate or strongly overestimate the span of time. Hence, most subjects have an awareness of the phenomenon of exponential growth bias. Despite this awareness, subjects exhibit exponential growth bias and mitigation bias in the frames using an exponential perspective.

## CONCLUSION

In the commonly used frame of case growth and daily exponential growth rates, subjects in the experiment drastically underestimate the benefit of decreasing the growth rate of an infectious disease. Such biased beliefs about the exponential spread of COVID-19 decrease the willingness to adhere to NPIs (*4*). We find that communicating exponential growth in terms of doubling times rather than growth rates decreases bias. Employing a logarithmic perspective, that is asking for time gained, rather than an exponential perspective, which asks for cases avoided, is even more effective at decreasing bias. The frame which combines doubling times with the logarithmic perspective fully eliminates mitigation bias. These findings suggest directions for changes that public health authorities could try to better communicate the benefits of NPIs and to increase adherence to them. Beyond public health, our findings may have applications to the regulation of the sale of financial products, retirement savings, education and the public understanding of exponential processes in the environment.

## MATERIALS AND METHODS

### Experimental Design

#### Experimental procedure

The online experiment was implemented by the laboratory staff of the ETH Decision Science Lab (DeSciL). The authors had no contact with the subjects, by e-mail or otherwise.

#### Treatment groups/frames

Subjects are randomly assigned with equal probability to one of the four treatment groups/frames. Each subject sees three questions related to exponential processes, each presented on a separate screen: the mitigation question and the high and low exponential growth questions. The question differ according to the treatment group/frame the subject is in. The question on beliefs about others’ answers also differs by treatment group/frame.

#### Questions and instructions

For all questions, the answer box is free form entry. No unit was specified. Subjects were instructed to always specify units in their answers wherever appropriate. Neutral examples on how to answer were used (for instance do not answer 187, answer 1 m 87 cm or answer 1.87 m).

Subjects were requested not to use calculators, spreadsheets or other tools. Rather, subjects were requested to use their intuition.

In the following we give the mitigation question, the high growth exponential question and the low growth exponential question. Brackets indicate the parts of the questions that differ across frames.

#### Mitigation question

In a country, 974 people have been infected so far. The number of infected people [grows by 26% daily]/[doubles every 3 days].

The country aims to have as few infected people as possible in 30 days. Therefore, the adoption of measures such as increased hand-washing and social distancing is being discussed. With these measures, the number of infected people would [grow at merely 9% per day]/ [double only every 8 days].

[How much time would the country gain with these measures until 1’000’000 people are infected?]/[How many infections could be avoided in the following 30 days with these measures?].

#### High exponential growth question

In a country, 974 people have been infected so far. The number of infected people [grows by 26% per day]/[doubles every 3 days]. [How long will it take until 1’000’000 people are infected in this country?]/[How many people will be infected in 30 days?]

#### Low exponential growth question

In a country, 974 people have been infected so far. The number of infected people [grows by 9% per day]/[doubles every 8 days]. [How long will it take until 1’000’000 people are infected in this country?]/[How many people will be infected in 30 days?]

A remark on the screen clarifies that in answering people who have died or recovered are to be taken into account as well.

#### Randomization of screen order

Questions are randomized such that subjects either see the screen with the mitigation question first, or the two exponential growth questions first. The two exponential growth questions were further randomized within each other. That is, within each frame subjects were randomly assigned to one of four subgroups, which saw the exponential questions in one of the following orders:

- High exponential growth question, low exponential growth question, mitigation question
- Low exponential growth question, high exponential growth question, mitigation question
- Mitigation question, high exponential growth question, low exponential growth question
- Mitigation question, high exponential growth question, low exponential growth question

#### Awareness of exponential growth bias

To elicit whether a subject is aware of the phenomenon of exponential growth bias, the high exponential growth question of their frame is again shown to the subject. She is asked to indicate her belief of how other subjects answered this question on a 5-point-Likert scale with the following options:

- Frames C-r and C-d: The answers of most participants were far too low / The answers of most participants were too low / The answers of most participants were approximately correct / The answers of most participants were too high / The answers of most participants were far too high.
- Frames T-r and T-d: Most participants indicated a timespan which was far too short / Most participants indicated a timespan which was too short / Most participants were approximately correct / Most participants indicated a timespan which was too long / Most participants indicated a timespan which was far too long.

### Statistical Analysis

#### Fraction of biased subjects

To test whether the fraction of biased subjects is larger in one frame than another, we convert the answers to 1 if the answer is below the true value, 0 otherwise. We then test the hypothesis that the probability of success is larger in the first frame than the second using Pearson’s chi-squared test.

#### Median answer

To test whether the median answer differs between two frames, we use the Brown-Mood median test.

#### Relation of biases

To test whether mitigation bias can be fully explained by exponential growth bias, we use a binomial test: Define as the number of successes the number of subjects who give an answer to the mitigation question which is larger than the difference between their answers to the exponential growth questions. Define as the number of failures the number of subjects who give an answer to the mitigation question which is smaller than the difference between their answers to the exponential growth questions.

## Data Availability

Data is available from the authors. We are uploading the associated data file (Excel) to medrxiv.org.

## General

We thank E. Ash, S. Bechtold G. Gertsch, and F.H. Schneider for insightful comments.

## Author contributions

M.S. and D.S. contributed equally to the project and are listed alphabetically.

## Supplementary Materials

**Fig S1.**
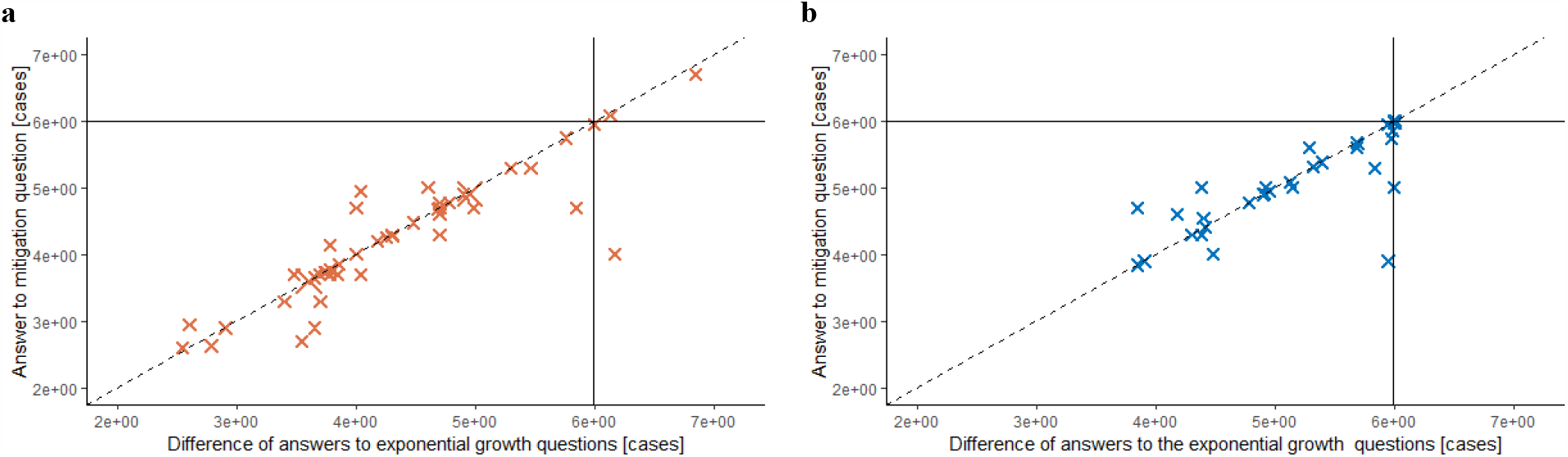
Mitigation bias and exponential growth bias – subsample of subjects who see exponential growth questions first, **a**, answers to the mitigation question plotted against the difference in answers to the exponential growth questions for frame C-r (n=49), **b**, same plot for frame C-d (n=40). Solid line indicates the correct answer (about 1 million cases avoided). For observations on the dashed line, mitigation bias can be fully explained by exponential growth bias (28% in frame C-r, 23% in frame C-d). Multiple identical answers displayed by larger crosses. Only subjects to whom the two exponential growth questions was displayed prior to the mitigation question. Data points with non-positive values are excluded.

**Table S1.** Question parameters and response quantiles

| Question/<br>Frame | Question parameters |  |  |  |  | Correct answer |  | Response quantiles |  |  |  |  |
| --- | --- | --- | --- | --- | --- | --- | --- | --- | --- | --- | --- | --- |
|  | Initial cases | Growth rate | Doubling time [days] | Period [days] | Final cases | Cases | Days | 1% | 25% | 50% | 75% | 99% |
| Mitigation question |  |  |  |  |  |  |  |  |  |  |  |  |
| C-r | 974 | 26% / 9% | - | 30 | - | 986,330 | - | 18.69 | 4,000 | 8,600 | 50,000 | 4,506,000 |
| C-d | 974 | - | 3 / 8 | 30 | - | 984,271 | - | 349 | 10,000 | 82,000 | 745,000 | 9,542,800 |
| T-r | 974 | 26% / 9% | - | - | 1,000,000 | - | 50.46 | 5.16 | 28 | 60 | 120 | 1,087.4 |
| T-d | 974 | - | 3 / 8 | - | 1,000,000 | - | 50.02 | 8.1 | 30 | 50 | 70 | 180 |
| High exponential growth question |  |  |  |  |  |  |  |  |  |  |  |  |
| C-r | 974 | 26% | - | 30 | - | 999,253 | - | 814 | 8,000 | 15,000 | 100,000 | 10,000,000 |
| C-d | 974 | - | 3 | 30 | - | 997,376 | - | 3,090 | 42,500 | 256,000 | 1,000,000 | 14,500,000 |
| T-r | 974 | 26% | - | - | 1,000,000 | - | 30.00 | 4 | 14 | 30 | 60 | 3,519.6 |
| T-d | 974 | - | 3 | - | 1,000,000 | - | 30.01 | 6.1 | 20 | 30 | 30 | 405.5 |
| Low growth exponential growth question |  |  |  |  |  |  |  |  |  |  |  |  |
| C-r | 974 | 9% | - | 30 | - | 12,923 | - | 93.2 | 2,907.5 | 5,000 | 30,000 | 10,000,000 |
| C-d | 974 | - | 8 | 30 | - | 13,105 | - | 2,550 | 8,000 | 15,000 | 20,000 | 1,000,000 |
| T-r | 974 | 9% | - | - | 1,000,000 | - | 80.46 | 7.13 | 35 | 90 | 172.5 | 1,400.2 |
| T-d | 974 | - | 8 | - | 1,000,000 | - | 80.03 | 10 | 50 | 80 | 88 | 802.75 |

## References and Notes

1. Lai, S., Ruktanonchai, N.W., Zhou, L. et al. Effect of non-pharmaceutical interventions to contain COVID-19 in China. Nature (2020). https://doi.org/10.1038/s41586-020-2293-x

2. Flaxman, S., Mishra, S., Gandy, A. et al. Estimating the effects of non-pharmaceutical interventions on COVID-19 in Europe. Nature (2020). https://doi.org/10.1038/s41586-020-2405-7

3. Hsiang, S., Allen, D., Annan-Phan, S. et al. The effect of large-scale anti-contagion policies on the COVID-19 pandemic. Nature (2020). https://doi.org/10.1038/s41586-020-2404-8

4. Lammers, J., Crusius, J. & Gast, A. Correcting misperceptions of exponential coronavirus growth increases support for social distancing. Proceedings of the National Academy of Sciences 117 (28), 16264–16266 (2020). https://doi.org/10.1073/pnas.2006048117

5. Wagenaar, W.A. & Timmers, H. The pond-and-duckweed problem: Three experiments on the misperception of exponential growth. Acta Psychologica 43, 239–251(1979).

6. Wagenaar, W.A. & Sagaria, S.D. Misperception of exponential growth. Perception and Psychophysics 18, 416–422 (1975).

7. Wagenaar W.A., Timmers H. (1978) Intuitive Prediction of Growth. In: Burkhardt D.F., Ittelson W.H. (eds) Environmental Assessment of Socioeconomic Systems. NATO Conference Series, vol 3. Springer, Boston, MA.

8. Christandl, F. & Fetchenhauer, D. How laypeople and experts misperceive the effect of economic growth. Journal of Economic Psychology 20, 381–392 (2009).

9. McKenzie, C.R.M. & Liersch, M.J. Misunderstanding savings growth: Implications for retirement savings behavior. Journal of Marketing Research 48, 1–13 (2011).

10. Almenberg, J. & Gerdes, C. Exponential growth bias and financial literacy. Applied Economic Letters 19, 1693–1696 (2012).

11. Goda, G. S., Flaherty Manchester, C. & Sojourner, A. J. What will my account really be worth? Experimental evidence on how retirement income projections affect saving. Journal of Public Economics 119, 80–92 (2014).

12. Levy, M. R. & Tasoff, J. Exponential growth bias and lifecycle consumption. Journal of the European Economic Association 14, 545–583 (2016).

13. Ambuehl, S., Bernheim, B. D. & Lusardi A. A method for evaluating the quality of financial decision making with an application to financial education. National Bureau of Economic Research working paper 20618 (2017).

14. Levy, M. R. & Tasoff, J. Exponential-growth bias and overconfidence. Journal of Economic Psychology 58, 1–14 (2017).

15. Bazerman, M.H. The Relevance of Kahneman and Tversky’s concept of framing to organizational behavior, Journal of Management, 10 (3), 333–343 (1984).

16. Camerer, C., Thaler, R. Anomalies: Ultimatums, Dictators and Manners, Journal of Economic Perspectives, 9 (2), 209–219 (1995).

17. Savage, L.J. The Foundations of Statistics, New York, Wiley (1954) 101–104.

18. Raiffa, H. Decision Analysis: Introductory Lecturs on Choices under Uncertainty. Reading, Mass. Addison-Wesley. (1968) 80–86.

19. Kahneman, D. & Tversky, A. An Analysis of decision under risk. Econometrica 47(2), 263–291 (1979).

20. Hershey, J.C. & Schoemaker, P.J.H. Risk taking and problem context in the domain of losses: an expected utility analysis. Journal of Risk and Insurance 47 (1), 111–132 (1980).

21. Tversky, A. & Kahneman, D. The framing of decisions and the psychology of choice. Science 211 (4481), 453–458 (1981).

22. McNeil, B.J., Pauker, S.G., Sox, H.C. & Tversky, A. On the elicitation of preference for alternative therapies. New England Journal of Medicine 306, 1259–1262 (1982).

23. Tversky, A. & Kahneman, D. Rational choice and the framing of decisions. Journal of Business 59 (4), S251–S278 (1986).

24. Howard, K. & Salkeld G. Does attribute framing in discrete choice experiments influence willingness to pay? Results from a discrete choice experiment in screening for colorectal cancer. Value in Health 12 (2), 354–363 (2009).

25. Dehaene, S. Symbols and quantities in parietal cortex: elements of a mathematical theory of number representation and manipulation. In Sensorimotor Foundations of Higher Cognition, Haggard, P., Rossetti, Y. & Kawato, M. (eds.), 527–574 (2008).

26. Dehaene, S., Izard, V. Spelke, E. & Pica, P. Log or linear? Distinct intuitions of the number scale in Western and Amazonian indigene cultures, Science, 320 (5880), 1217–1220 (2008).

27. Khāllikan, I. Biographical Dictionary (MacGuckind de Slane, W. Oriental Translation Fund of Great Britain and Ireland, 1868), vol. 3 (1274).

28. Alighieri, D. “Canto XXVIII” in Paradiso, line 93 (early 14th century).

29. Zeit. Schach und das Virus: Warum das Ausbreitungstempo bei Corona so entscheidend ist. Die Zeit. https://www.zeit.de/news/2020-03/12/warum-das-ausbreitungstempo-bei-corona-so-entscheidend-ist (12 March 2020).

30. Dalton, C. I’m an ER doctor. Please take coronavirus seriously. The Guardian. https://www.theguardian.com/commentisfree/2020/mar/20/er-doctor-coronavirus-exponential-growth (20 March 2020).

